# Scale-up of 99DOTS for tuberculosis treatment supervision in Uganda: An interrupted time series analysis

**DOI:** 10.1101/2024.01.22.24300949

**Authors:** Rebecca Crowder, Suzan Nakasendwa, Alex Kityamuwesi, Muhammad Musoke, Joyce Nannozi, Joseph Waswa, Agnes Nakate Sanyu, Maureen Lamunu, Amon Twinamasiko, Lynn Kunihira Tinka, Denis Oyuku, Diana Babirye, Christopher Berger, Ryan Thompson, Stavia Turyahabwe, David Dowdy, Achilles Katamba, Adithya Cattamanchi, Noah Kiwanuka

**Affiliations:** Division of Pulmonary and Critical Care Medicine, San Francisco General Hospital, University of California San Francisco, San Francisco, USA; Center for Tuberculosis, University of California San Francisco, San Francisco, USA; Uganda Tuberculosis Implementation Research Consortium, Kampala, Uganda; Department of Epidemiology, Johns Hopkins Bloomberg School of Public Health, Baltimore, USA; National Tuberculosis and Leprosy Program, Uganda Ministry of Health, Kampala, Uganda; Clinical Epidemiology & Biostatistics Unit, Department of Medicine, Makerere University College of Health Sciences, Kampala, Uganda; Division of Pulmonary Diseases and Critical Care Medicine, University of California Irvine, Irvine, USA; Department of Epidemiology and Biostatistics, School of Public Health, Makerere University College of Health Sciences, Kampala, Uganda

**Keywords:** digital adherence technology

## Abstract

**Background:** Digital adherence technologies like 99DOTS are being scaled-up for tuberculosis (TB) treatment despite limited evidence of their effectiveness and concerns about accessibility. We evaluated the reach and effectiveness of 99DOTS during its scale-up in Uganda.

**Methods:** We included all adults initiating drug-susceptible pulmonary TB treatment between August 2019 and June 2021 at 12 99DOTS-naïve health facilities (n=3,526). Using an interrupted time series design, we compared the proportion with treatment success (primary outcome) and enrolled on 99DOTS in the 9 months before and 12 months after implementing an ‘enhanced 99DOTS’ intervention. This included providing low-cost phones to people with TB when needed, task shifting monitoring and support to community health workers, and automated task lists. Treatment data were derived from routine TB treatment registers.

**Results:** The proportion enrolled on 99DOTS post-intervention was high (87.2%) and had a slight upward trend (proportion ratio 1.01, 95% CI 1.00-1.01). Treatment completion remained similar across periods (89.8% post-intervention vs. 87.1% pre-intervention). There was no immediate level or slope change in treatment success following the intervention.

**Conclusions:** With appropriate implementation supports, 99DOTS can have high uptake without negatively impacting treatment outcomes. Equity in access should be prioritized during implementation

---

Suboptimal tuberculosis (TB) treatment outcomes remain a major challenge in high TB burden countries. Digital adherence technologies (DATs) have been proposed as one potential intervention to support people with TB to successfully complete treatment. 99DOTS is a low-cost DAT, whereby people with TB self-report medication dosing by calling toll-free phone numbers hidden underneath pills in blister packs.^1, 2^ 99DOTS has been deployed in some Asian and African countries, including India,^1, 3^ Uganda ^2^ and Kenya.^4^

DATs are being scaled-up despite limited evidence of their effectiveness and concerns about accessibility to the technologies. In the only randomized trial of 99DOTS to date, we showed that only 52% of people with TB were enrolled on 99DOTS, and 99DOTS-based treatment supervision did not improve treatment outcomes.^2^ Other implementation studies have found similarly low enrollment of people with TB on 99DOTS and have reported similar treatment outcomes between routine- and 99DOTS-based treatment supervision.^1-3^

Barriers to the use of 99DOTS are now better documented. With respect to reach, consistent access to a phone has been reported as a key barrier among people with TB in high burden countries.^5^ It is unclear whether people without phone access would benefit from 99DOTS if this barrier were removed. In addition, the potential impact of 99DOTS is limited if health workers are unable to use daily dosing data to enhance treatment monitoring and support.^6^ Factors impacting health worker engagement with 99DOTS include technology literacy,^5^ usability of the 99DOTS application, and demanding workloads.^6^

To address these factors, we developed an implementation strategy for 99DOTS-based treatment supervision (‘enhanced 99DOTS’) that included components to increase uptake (providing low-cost phones to people with TB when needed) and enhance treatment monitoring and support (task shifting to community health workers and automated task lists). In this study, we aimed to assess whether the enhanced 99DOTS strategy improved uptake of 99DOTS and TB treatment outcomes among adults with pulmonary TB (PTB).

## METHODS

### Study design and population

We conducted an interrupted time series (ITS) analysis to evaluate the effect of the enhanced 99DOTS-strategy at 12 health facilities that did not participate in the implementation trial (*i*.*e*., 99DOTS-naïve facilities). These facilities included 8 Level III and 4 Level IV Health Centers in the Greater Kampala Metropolitan Area (Greater Kampala). These facilities were selected because they 1) treated >10 people with PTB/month in 2017 (sufficient to assess outcomes), 2) had a PTB treatment success rate in 2017 <80% (able to show impact), and 3) were classified as a lower-level health facility (i.e., Health Center III or IV). Of the 139 TB treatment units in Greater Kampala, these 12 met the eligibility criteria and were all selected in consultation with the Uganda National TB and Leprosy Programme (NTLP).

We included all adults initiating treatment for drug susceptible PTB at these facilities between August 1, 2019, and June 30, 2021. We excluded people with TB who were transferred out to another facility during treatment, as they would not have had the opportunity to fully benefit from the intervention.

This study was approved by institutional review boards at Makerere University School of Public Health and the University of California San Francisco, and by the Uganda National Council for Science and Technology. A waiver of informed consent was granted to access the demographic and clinical information recorded in TB treatment registers.

### Intervention periods

During the 9-month pre-intervention period (August 2019-April 2020), routine TB treatment support and supervision was provided via community-based DOT. The implementation of community-based DOT varied by health facility.^7^

During the buffer period (May-June 2020), study staff conducted training on the enhanced 99DOTS strategy via 2-to 3-day site visits at all participating health facilities.

During the subsequent 12-month post-intervention period (July 2020-June 2021), the enhanced 99DOTS strategy was available for TB treatment support and supervision at all participating health facilities (**Supplemental Figure 1**). The enhanced 99DOTS strategy included all components of the original 99DOTS intervention (automated dosing reminders via SMS and dosing confirmation via toll free calls as described for the DOT to DAT trial).^8, 9^ In addition, based on lessons from the trial, we 1) provided low-cost phones ($8 USD) to people with TB who lacked a phone or did not have regular access to a phone; 2) task shifted adherence monitoring and follow-up from clinicians to community health workers; and 3) provided automated task lists to facilitate community health worker follow-up of dosing history.

### Procedures

Data on TB treatment initiation and outcomes, as well as demographic and clinical characteristics, were collected from routine TB treatment registers at each health facility. Facility staff were trained to take and upload photos of the register to a secure, password protected server. Research staff then extracted data from these photos into a secure database using Research Electronic Data Capture (REDCap) software.^10^ Enrollment onto 99DOTS was confirmed by merging this database with a list of enrolled participants extracted from the 99DOTS server. Enrollment on 99DOTS was defined as enrollment within the first month (28 days) of TB treatment.

### Outcomes

The primary outcome was the proportion of people with TB treated successfully, defined as being assigned a treatment outcome of cured or completed in the Unit TB Treatment register. Secondary outcomes included the proportion of people with TB enrolled on 99DOTS for TB treatment support and supervision, the proportion persisting on treatment through the intensive phase (defined as completing 56 doses of treatment) and the proportion lost to follow-up.

### Statistical analysis

Interrupted time series analysis was used to model the change in mean proportions of TB treatment outcomes and reach of 99DOTS before and after the implementation of the enhanced 99DOTS-based intervention. We assessed for immediate (level) changes in outcomes, as well as changes in trend (slope).

The primary and secondary outcomes were analyzed according to intention-to-treat (ITT) and per protocol (PP) principles. The ITT analysis included all eligible participants, whereas the PP analysis excluded those who did not receive the intended method of TB treatment supervision, including pre-intervention participants who later received 99DOTS and post-intervention participants who never received 99DOTS or received 99DOTS late into their treatment (>28 days after treatment start).

Trajectory plots of observed and predicted outcomes were inspected to assess the overall post intervention linearity. Single group ITS models were fit using the mean proportion with each outcome of interest by health facility-month (see **Supplemental Methods in Appendix** for equation) using the Stata command *itsa*.^11^ Autocorrelation was assessed using the Cumby-Huizinga test and adjusted for as needed in the final models. We concluded that serial autocorrelation was present when the p-value for three consecutive lags (up to 21 lags tested) was significant (p<0.05). If there was borderline evidence of autocorrelation, a sensitivity analysis was performed, and the later lag was only used if standard errors of all parameters of interest were smaller. Proportion ratios (PR) were output from the ITS model to characterize pre- and post-intervention change in level (immediate change in outcome at time of enhanced 99DOTS implementation) and change in slope (average change in outcome per month). Stata version 18 was used for all analyses.

## RESULTS

### Study Population

From August 1, 2019, to June 31, 2021, 4,253 adults were initiated on treatment for drug-susceptible PTB at the 12 participating health facilities. The ITT population included 3,526 eligible people with TB (**Figure 1**), and the PP population included 3,197 people with TB. The PP population excluded people with TB who did not receive the intended intervention (standard of care during pre-intervention period or 99DOTS during the post-intervention period).

**Figure 1.**
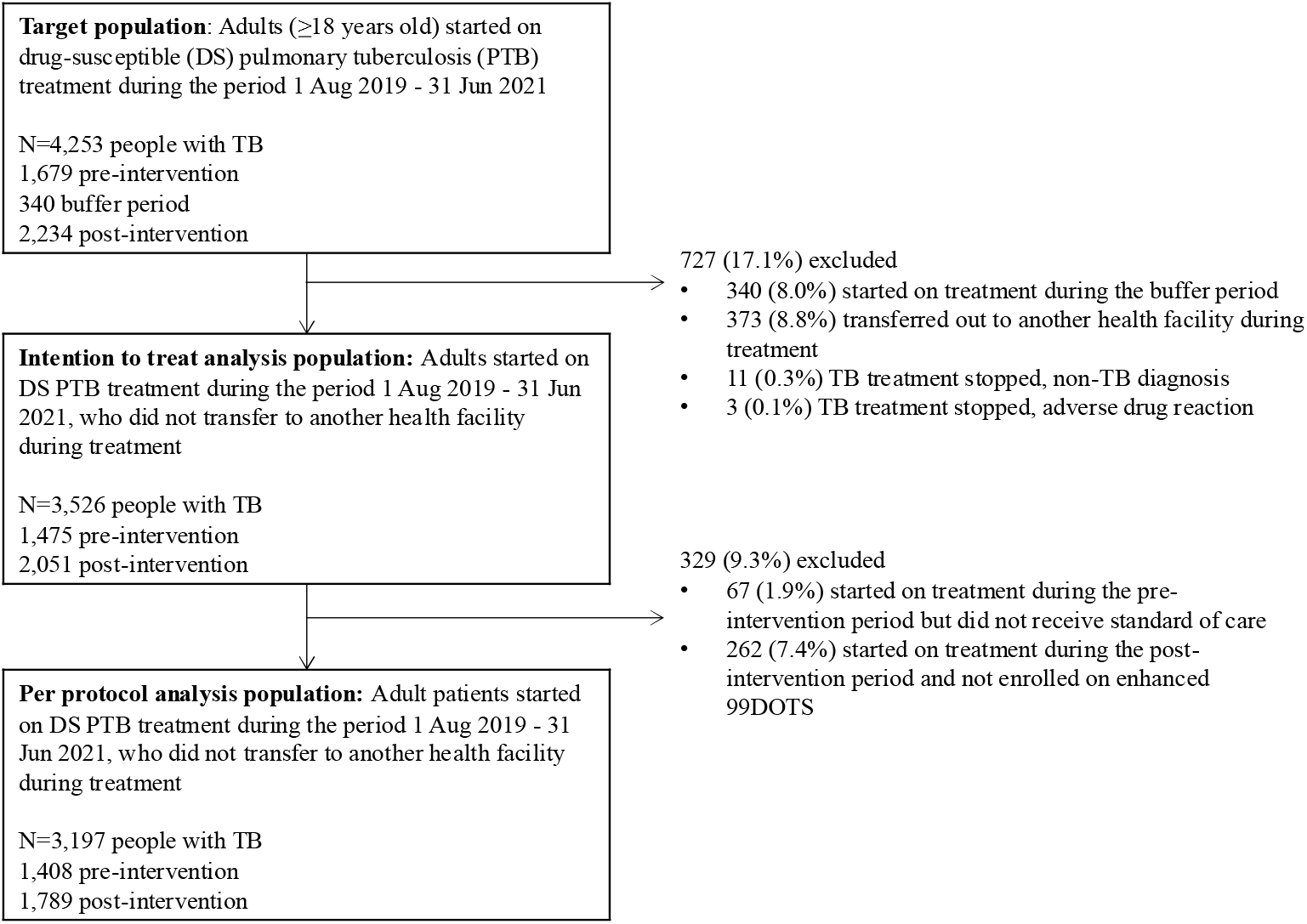
Target and analysis population.

There were no differences between the pre- and post-intervention period ITT populations in the proportions female, living with HIV, receiving ART (if HIV-positive), or with bacteriologically-confirmed TB. However, median age was higher (35 vs. 33, p<0.001) and the proportion with previous TB lower (6.5% vs. 8.5%, p=0.02) in the post-intervention period (**Table 1**).

**Table 1.** Participant baseline characteristics by study period and facility type.

|  | Intention-to-treat population |  |  | Per protocol population |  |  |
| --- | --- | --- | --- | --- | --- | --- |
|  | Pre-intervention Period<br>(n = 1,475) | Post-Intervention Period<br>(n = 2,051) | P-value* | Pre-intervention Period<br>(n = 1,408) | Post-Intervention Period<br>(n = 1,789) | P-value* |
| Age in years, median (IQR) | 33 (26-42) | 35 (28-43) | <0.001 | 33 (26-42) | 35 (28-43) | <0.001 |
| Female, n (%) | 504 (34.2) | 711 (34.7) | 0.76 | 478 (34.0) | 619 (34.6) | 0.70 |
| HIV positive, n (%) | 662 (44.9) | 897 (43.7) | 0.50 | 635 (45.1) | 795 (44.4) | 0.71 |
| On ART, n (%) | 662 (100) | 897 (100) | - | 635 (100) | 795 (100) | - |
| Previous TB, n (%) | 126 (8.5) | 133 (6.5) | 0.02 | 116 (8.2) | 101 (5.7) | 0.004 |
| Bacteriologically confirmed TB, n (%) | 867 (58.8) | 1,265 (61.7) | 0.08 | 828 (58.8) | 1,117 (62.4) | 0.04 |

\* Continuous variables were compared using a Wilcoxon rank sum test, and categorical variables were compared using a chi-square test.

Comparisons in the PP population were similar, except that there were more people with bacteriologically confirmed TB in the post-intervention period (62.4% vs. 58.8% in the pre-intervention period, p=0.04).

There were no missing outcome data in the key variables reported.

### Enrollment on 99DOTS

At these 99DOTS-naive facilities, 99DOTS was not offered during the pre-intervention period. Enrollment on 99DOTS was immediately high (n=1,789, 87.2%) following implementation of the enhanced 99DOTS strategy and had an upward trend during the post-intervention period (PR 1.01, 95% CI 1.00-1.01) (**Figure 2, Table 2)**.

**Table 2.** Interrupted time series regression results from an unadjusted model.

|  | N | Change in level<br>(PR, 95% CI) | Change in slope<br>per month<br>(PR, 95% CI) | Pre-intervention<br>slope per month<br>(PR, 95% CI) | Post-intervention<br>slope per month<br>(PR, 95% CI) |
| --- | --- | --- | --- | --- | --- |
| Reach: Enrolled onto 99DOTS |  |  |  |  |  |
| ITT | 3,526 | - | - | - | 1.01 (1.00, 1.01)* |
| Treated successfully |  |  |  |  |  |
| ITT | 3,526 | 0.99 (0.94, 1.05) | 0.99 (0.98, 1.00) | 1.01 (1.00, 1.02) | 1.00 (1.00, 1.00) |
| PP | 3,197 | 1.04 (0.95, 1.14) | 0.99 (0.98, 1.00) | 1.01 (0.99, 1.02) | 1.00 (0.99, 1.00) |
| Completion of the intensive phase |  |  |  |  |  |
| ITT | 3,526 | 0.98 (0.92, 1.04) | 0.99 (0.98, 1.00) | 1.01 (1.00, 1.02)^ | 1.00 (1.00, 1.00) |
| PP | 3,197 | 1.02 (0.94, 1.10) | 0.99 (0.98, 1.00) | 1.01 (1.00, 1.02) | 1.00 (1.00, 1.00) |
| Lost to follow-up |  |  |  |  |  |
| ITT | 3,526 | 1.04 (1.01, 1.06)* | 1.01 (1.00, 1.01) | 0.99 (0.98, 1.00)* | 1.00 (0.99, 1.00)* |
| PP | 3,197 | 1.00 (0.96, 1.05) | 1.01 (1.00, 1.01) | 0.99 (0.98, 1.00) | 1.00 (0.99, 1.00) |

PR: Proportion Ratio; ITT: intention to treat; PP: per protocol
Statistically significant results noted with \* ( $p < 0.05$ ), ^ ( $0.05 < p < 0.06$ , borderline)

**Figure 2.**
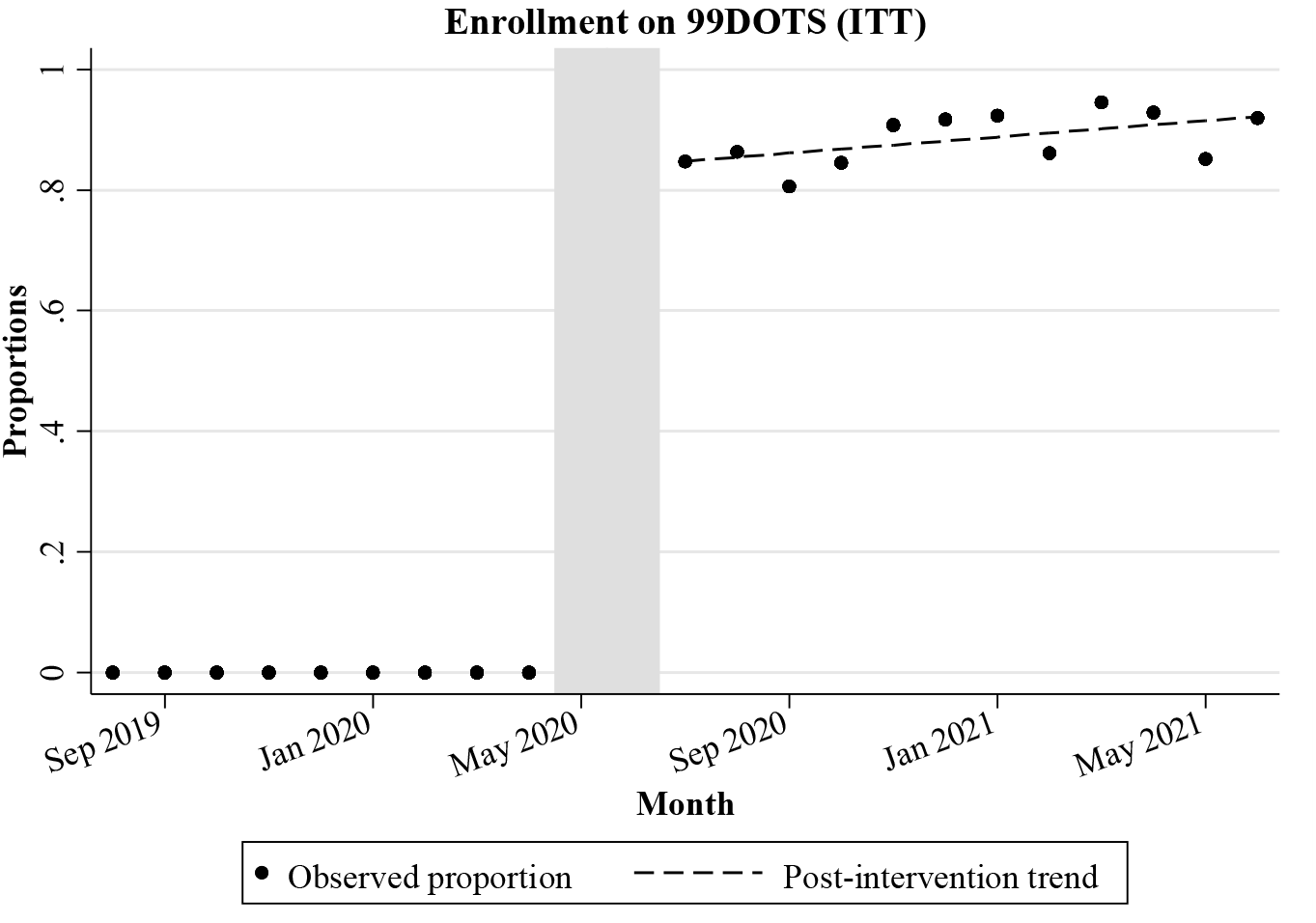
Enrollment on 99DOTS. This graph presents the proportion of adults initiating PTB treatment at 12 participating health facilities that were enrolled on 99DOTS each month. Health worker training on the 99DOTS intervention was conducted between May-June 2020, after which the enhanced 99DOTS strategy was available for TB treatment supervision. The proportion enrolled on 99DOTS post-intervention was high (87.2%) and had a slight upward trend (proportion ratio 1.01, 95% CI 1.00-1.01).

### Treatment success (primary outcome)

#### Intention-to-treat analysis

Treatment outcomes largely remained stable over the pre- and post-intervention periods. 1,285 (87.1%) people with TB successfully completed treatment during the pre-intervention period, and 1,841 (89.8%) successfully completed treatment during the post-intervention period. The ITS analysis showed no significant change in level or slope of treatment success **(Figure 3, Table 2)**.

**Figure 3.**
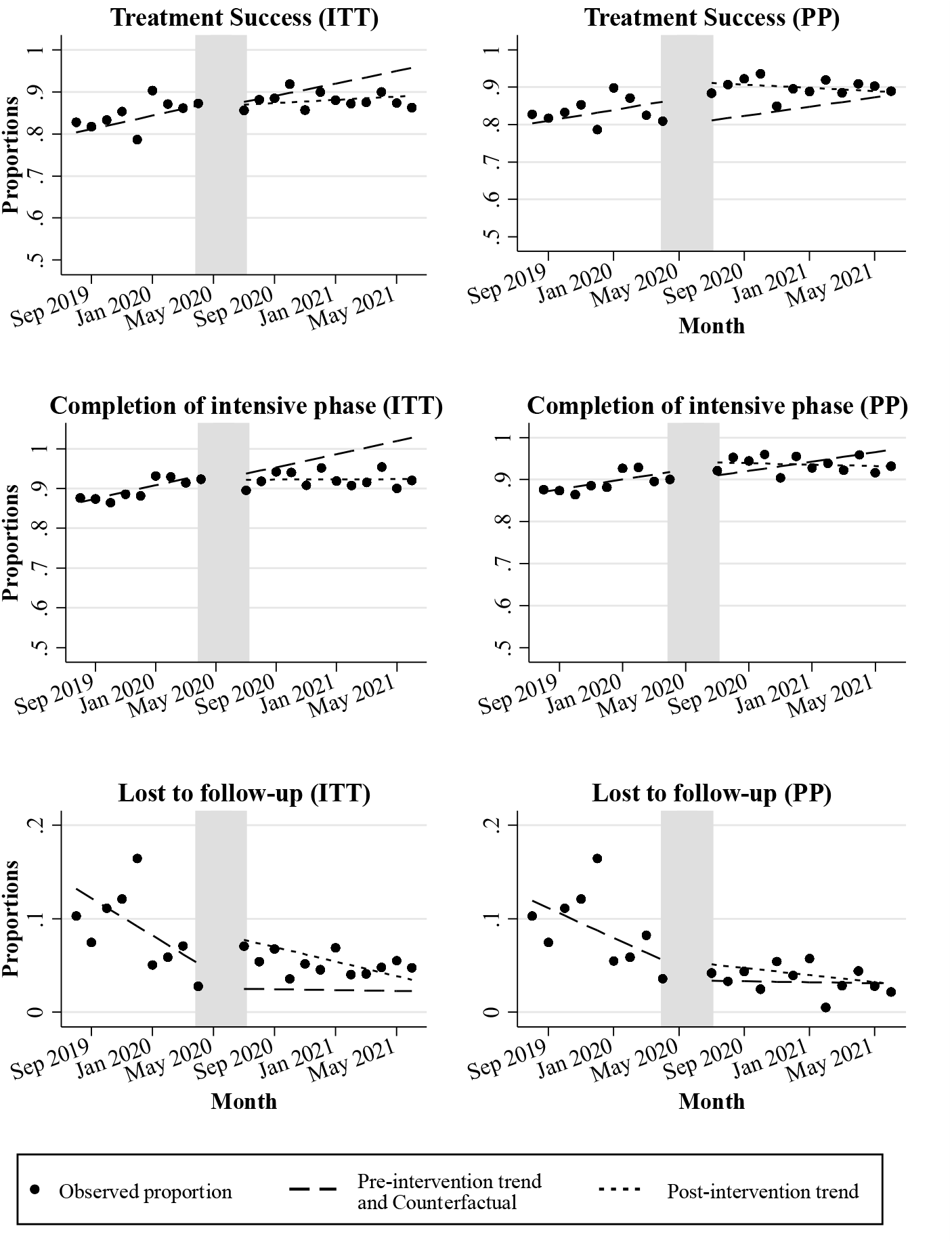
Interrupted time series analysis of TB treatment outcomes before and after implementation of enhanced 99DOTS for TB treatment supervision. The long, dashed lines present the expected post-intervention trends had the pre-intervention trends continued. For comparison, the short, dashed lines show the observed post-intervention trends. ITT: intention to treat; PP: per protocol

#### Per-protocol analysis

Similarly, in the per protocol analysis, treatment success remained stable during the pre- and post-intervention periods; 1,219 (86.6%) people with TB successfully completed treatment during the pre-intervention period and 1,629 (91.1%) successfully completed treatment during the post-intervention period. There was no significant change in level or slope of treatment success (**Figure 3, Table 2**).

### Other secondary outcomes

#### Completion of intensive phase

The proportion completing the intensive phase of treatment increased from 1,353 (91.7%) during the pre-intervention period to 1,917 (93.5%) during the post-intervention period; however, there was no significant change in level or slope following the intervention **(Figure 3, Table 2**).

#### Loss to follow-up

The proportion lost to follow-up during the post-intervention period was lower than during the pre-intervention period (91 [4.4%] vs. 108 [7.3%]). Loss to follow-up was decreasing before and after the intervention. There was a significant level increase in loss to follow-up following implementation of enhanced 99DOTS and no change in slope (negative during both periods, **Figure 3, Table 2**).

## DISCUSSION

DATs are increasingly being considered as a more person-centered alternative to directly observed therapy for TB treatment supervision and support. In this quasi-experimental study of an enhanced 99DOTS strategy, we found that 87% of adults with PTB were enrolled on 99DOTS and achieved similar treatment outcomes compared to routine care prior to 99DOTS implementation. These findings suggest that 99DOTS can eliminate the need for directly observed therapy for most people with TB without compromising treatment outcomes.

Low enrollment has been a challenge in previous studies of 99DOTS and other cellphone-based DATs, raising concerns about equity and access to this mode of care. Access to mobile phones is expected to be lower among people with TB than in the general population (18% vs. 7.8% in Peru from 2007-2013).^12^ Mobile phone access among populations affected by TB in Uganda has been reported to be high (89% among PLHIV with latent TB in a cohort in western Uganda^13^ and 58% ownership with an additional 42% indirect access among household contacts of people with TB in urban Kampala.^14^ Nevertheless, an intervention that relies on cellphone access systematically excludes some of the most vulnerable. Provision of low-cost phones in this study significantly improved the reach of 99DOTS compared to our previous trial in Uganda.^2^ This demonstrates the feasibility of a critical component for equity of any digital intervention requiring a cellphone.

Most studies evaluating the impact of DATs on treatment outcomes have shown similar or negative effects. A systematic review in 2022 of 16 RCTs evaluating DATs, including 10 that assessed impact on treatment completion, found that 4/10 reported a positive impact on treatment completion, 1/10 reported a significantly negative impact, and 5/10 reported no significant impact on treatment completion.^15^ The version of 99DOTS implemented in this study was enhanced using human-centered design methods, which offer benefits over SMS or phone reminders alone.^16, 17^ Despite these improvements and further refinement from the previous version of 99DOTS implemented in Uganda,^9^ our study found that treatment success rates were similar before and after the implementation of enhanced 99DOTS and remained below the WHO target of 90%. Cumulative evidence suggests DATs alone do not improve treatment outcomes.

While DATs can make TB treatment more convenient and less costly in some cases, they do not address structural factors associated with non-adherence such as poverty, gender discrimination, social influences, or the catastrophic costs incurred during TB treatment.^18^

A strength of this study was its pragmatic approach and generalizable population. We included all adults initiating treatment for drug-susceptible pulmonary TB across 12 health facilities in Uganda over almost two years. Together with the facilities included in the previous trial, this represents about 10% of the population with TB in Uganda. By offering phones to all people with TB who lacked access, we removed the barrier of phone access and the associated confounding hindering previous studies. Our study also has some limitations. ITS analyses can be impacted by time-varying confounders that do not remain consistent across the pre- and post-intervention periods. In this case, the study observation period included the beginning of the COVID-19 pandemic. In Uganda, shelter in place policies, including a ban on public transportation, went into effect at the end of March 2020. 99DOTS training was permitted to take place with appropriate precautions during visits to the health facilities in May and June 2020. Following the shelter-in-place order, TB case notifications in Uganda declined immediately,^19^ which was consistent with our data and may indicate a different population observed during the post-intervention period. If the population initiated on treatment after the shelter-in-place policies went into effect were sicker due to selection bias, we would expect them to have a lower odds of treatment success, thus biasing these results toward the null. Treatment outcomes, measured by proportion, remained stable after the policy change, allowing comparison before and after the 99DOTS intervention.

In conclusion, 99DOTS can enable treatment support and monitoring for people with TB without compromising treatment outcomes. However, further intervention to address the root causes of poor adherence will be needed to achieve END TB targets. 99DOTS and other DATs may still play a role in building a more person-centered TB care system, offering more flexibility over traditional directly observed therapy. Equity should be prioritized in implementation, including provision of low-cost phones when required.

## Data Availability

All data produced in the present study are available upon reasonable request to the authors.

## Acknowledgements

We thank the administration, staff, and patients at participating health facilities. The study was implemented in collaboration with the Uganda National TB and Leprosy Program and was made possible by the commitment of district TB focal persons and health facility staff.

