## Appendix for "Scale-up of 99DOTS for tuberculosis treatment supervision in Uganda: An interrupted time series analysis"

**Supplemental Methods. Equation for interrupted time series regression model.**

*Y_it_* = *β*_0_  + *β*_1_*T* + *β*_2_ X_t_+  *β*_3_(*T**X_t_) + ϵ_t_

Where:

- *Y_it_* is the aggregated outcome in a given health facility during each study month
- *T* is the time since the start of the study, X_t_ is the intervention period indicator variable coded 0 for pre-intervention and 1 for post-intervention, and T*X_t_ is an interaction term
  - *β*_0_ is the intercept of the outcome for the pre-intervention period,
  - *β*_1_ is the slope for the pre-intervention period,
  - *β*_2_ is the level change following the intervention, with (*β*_0_ + *β*_2_) representing the intercept of the outcome for the post-intervention period,
  - *β*_3_ is the difference in post-intervention and pre-intervention period slopes, with (*β*_1_ + *β*_3_) representing the slope of the post-intervention period

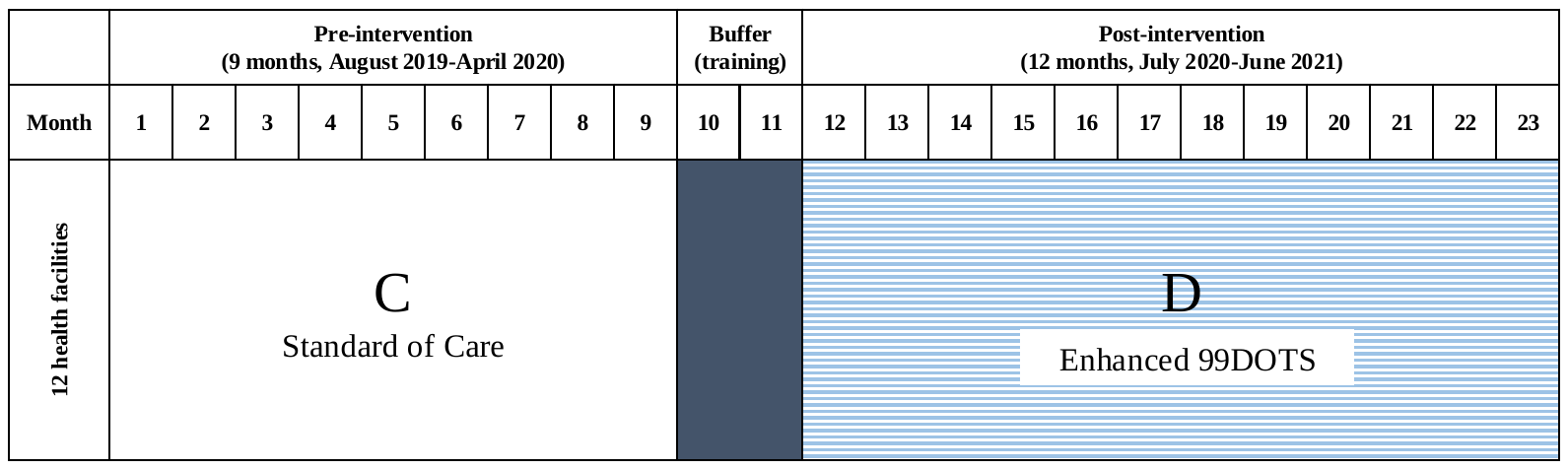

**Supplemental Figure 1. Interrupted time series study design**

**Supplemental Table 1. Proportion of treatment outcomes per site.**

| **99DOTS-naive sites (12 health facilities)** | | | | | | |
| --- | --- | --- | --- | --- | --- | --- |
|  | **Intention-to-treat population** | | | **Per protocol population** | | |
|  | **Pre-intervention Period (n = 1,475)** | **Post-Intervention Period (n = 2,051)** | **P-value*** | **Pre-intervention Period (n = 1,408)** | **Post-Intervention Period (n = 1,789)** | **P-value*** |
| **Reach: Enrolled onto 99DOTS** | - | 1,789 (87.2) | - | - | - | - |
| **Treated successfully** | 1,285 (87.1) | 1,841 (89.8) | 0.007 | 1,219 (86.6) | 1,629 (91.1) | 0.000 |
| **Completed intensive phase** | 1,353 (91.7) | 1,917 (93.5) | 0.025 | 1,286 (91.3) | 1,690 (94.5) | 0.000 |
| **Lost to follow-up** | 108 (7.3) | 91 (4.4) | 0.000 | 108 (7.7) | 57 (3.2) | 0.000 |

P-values obtained using proportion test
